# Nephroprotective Effect of GLP-1 Receptor Agonists (GLP-1 RAs) in Patients Receiving Lithium Therapy: A Population-Based Study Using the TriNetX Network

**DOI:** 10.64898/2026.02.09.26345925

**Authors:** Roger S. McIntyre, Yanli Zhang-James, Joseph F. Goldberg, Angela T.H. Kwan

## Abstract

GLP-1 receptor agonists (GLP-1 RAs) are effective in delaying progression of chronic kidney disease in individuals with type 2 diabetes mellitus (T2DM). We evaluated whether GLP-1 RA prescription is associated with reduced nephrotoxicity in adults receiving long-term lithium therapy. We conducted a retrospective, propensity score-matched cohort study using electronic health records from the TriNetX global network, which includes de-identified data from over 127 million patients across 109 healthcare organizations. The study population consisted of adults aged ≥18 years with T2DM, with lithium exposure within the 2 years preceding the index date and at least one prescription for a GLP-1 RA. The primary efficacy outcome was the rate of renal nephrotoxicity in persons with T2DM prescribed lithium and a GLP-1 RA versus those with T2DM prescribed lithium but no GLP-1 RA or other antidiabetic agents. Nephrotoxicity was a composite of ICD-10 and CPT-coded renal disease. Incidence and time-to-event outcomes were assessed using Kaplan-Meier curves and Cox proportional hazards models. In our 24-month analysis, 462 matched patient pairs were included. Initiation of a GLP-1 RA during lithium therapy was associated with a lower incidence of renal events versus lithium alone (6·1% vs 10·4%), corresponding to a risk difference of -4.3% (95% CI -7·86 to -0·80), a risk ratio of 0·58 (95% CI 0·37-0·91; p=0·017), and higher event-free survival (89·0% vs 83·2%; log-rank p=0·037). GLP-1 receptor agonist therapy was associated with a reduction in reports of lithium-associated nephrotoxicity. Our findings provide impetus to conduct mechanistic renal histopathologic studies combining GLP-1 RAs with lithium.

## Introduction

Lithium remains one of the most effective treatments for acute mania and long-term relapse prevention in bipolar I disorder, with benefits that extend beyond mood stabilization to include reduced suicide risk and possible neuroprotective effects.^1–3^ A major safety concern that requires regular laboratory monitoring and a common reason for both non-initiation and early discontinuation of lithium therapy is lithium-associated nephrotoxicity. Precisely estimating the frequency of lithium-associated chronic kidney disease in persons with long-term lithium exposure has been difficult because of multiple confounding factors that affect the risk estimate in the mood disorder population (e.g., type 2 diabetes mellitus [T2DM]).^4,5^ Nonetheless, the estimated population prevalence of chronic kidney disease (CKD) stage 3 or greater is 6-7%; in persons with T2DM, it is approximately 30-40%, while in persons with long-term lithium exposure, estimates have ranged from 27-32%.^6–8^ No intervention has been established as effective in the treatment or prevention of lithium-associated nephrotoxicity. Discontinuation of lithium in this context increases risk for the person with lived experience of bipolar disorder, and it has not necessarily been established that switching to an anticonvulsant (some of which are also associated with nephrotoxicity) consistently results in a reduction in chronic kidney disease progression.^4,9^ Neophrotoxicity due to lithium treatment has contributed to the reduction in use of lithium in individuals with bipolar disorder despite its longstanding position as the most effective maintenance treatment.^10,11^

GLP-1 receptor agonists (GLP-1 RAs) produce substantial glucose- and weight-lowering effects and are now widely used in the management of metabolic disease. GLP-1 receptor agonists also lower cardiovascular mortality in individuals with obesity; each of the aforementioned comorbidities are known to differentially affect persons living with depression or bipolar disorder.^12,13^ Pertinent to this analysis, GLP-1 RAs such as semaglutide are also FDA-approved to reduce the risk of worsening kidney disease and progression to kidney failure in adults with T2DM and chronic kidney disease.^14,15^

Mechanisms hypothesized as mediators of the renal protective effects of GLP-1 RAs in T2DM overlap with mechanistic pathways implicated in lithium-associated nephrotoxicity.^16^ For example, long-term lithium exposure alters intracellular signaling, influences cellular proliferation, promotes extracellular matrix deposition and fibrosis, and activates oxidative and inflammatory cascades.^17^ Further impetus to consider GLP-1 RAs as potential treatments and/or prevention strategies for persons receiving lithium is a separate line of evidence which has preliminarily reported the nephroprotective effects of GLP-1 RAs in the context of exposure to other nephrotoxic agents (e.g., cisplatin, tacrolimus).^18,19^ Whether GLP-1 RAs confer similar protection against lithium-associated nephrotoxicity has not been thoroughly evaluated in clinical populations.^20^ Herein, we examined whether GLP-1 RA exposure is associated with a reduction in a composite measure of lithium-related nephrotoxicity.

## Methods

### Study design and data source

We conducted a retrospective cohort study using the TriNetX Research Network, a federated cloud-based analytics platform that aggregates deidentified electronic health records from participating health systems worldwide.^21^ TriNetX harmonises demographic, diagnostic, procedural, medication, and laboratory data using the Observational Medical Outcomes Partnership (OMOP) common data model. All analyses were executed within the secure TriNetX Live environment, and no identifiable information was accessed. Small cell counts (n ≤ 10) were automatically rounded by TriNetX to comply with privacy rules. Institutional Review Board approval was obtained at SUNY Upstate Medical University (IRB #2302956-2).

### Study population

Eligible participants were adults aged 18 years or older with T2DM and at least one recorded lithium prescription. For each analytic window, individuals were required to have at least one lithium prescription during the 2 years preceding the index date of GLP-1 RA initiation and to have no GLP-1 RA exposure during that same period. Exclusion criteria included prior renal transplantation, active pregnancy, and missing demographic variables. The two year interval preceding the index date was selected as the primary exposure window to ensure that participants had sustained, multiyear lithium treatment before GLP-1 RA initiation.

GLP-1 receptor agonists exposure was defined using a prespecified concept set that included semaglutide, liraglutide, dulaglutide, exenatide, lixisenatide, and tirzepatide. Comparator patients were lithium-treated individuals who had no GLP-1 RA prescriptions during the 2 years before cohort entry and who remained unexposed to GLP-1 RAs during follow-up. For GLP-1 RA-exposed patients, the index date was defined as the date of the first qualifying GLP-1 RA prescription. For comparator patients, the index date was defined as the first qualifying lithium prescription. In both groups, exposure was required to occur within the same relative time window (from 24 months prior to the index date up to the index date). Each analytic window was constructed independently, and identical timing rules were applied to both groups.

### Comparator populations

It is established that GLP-1 RAs are nephroprotective in persons living with T2DM. Consequently, to explore whether GLP-1 RAs may confer protection against lithium-associated nephrotoxicity beyond their established renal benefits in T2DM, we conducted a sensitivity analysis comparing individuals with T2DM prescribed both a GLP-1 RA and lithium with those prescribed a GLP-1 RA alone. For both groups, the index date was defined as the first GLP-1 RA prescription, and kidney outcomes occurring before this date, including those arising after lithium initiation but prior to GLP-1 RA initiation, were excluded.

### Outcome measure and primary efficacy endpoint

The primary outcome was a composite renal endpoint defined using International Classification of Diseases and Current Procedural Terminology codes. Components included acute kidney failure (N17.x), acute tubular necrosis (N17.0), cortical necrosis (N17.1), medullary necrosis (N17.2), other acute kidney failure (N17.8), unspecified acute kidney failure (N17.9), unspecified renal failure (N19), dependence on dialysis (Z99.2), end-stage renal disease (N18.6), renal transplant complications (T86.10, T86.11, T86.12, T86.19), kidney transplant status (Z94.0), haemodialysis procedures (CPT 90935 and 90937), unspecified nephritic syndrome consistent with tubulointerstitial nephritis (N05.8), tubulointerstitial nephritis not specified as acute or chronic (N12), unspecified nephritic syndrome (N05), and other renal disorders (N28) when used clinically to indicate nephropathy.

The primary efficacy endpoint was the rate of composite nephrotoxicity among persons with T2DM prescribed both lithium and a GLP-1 RA compared with persons prescribed lithium but not a GLP-1 RA. Our sensitivity analysis evaluated persons prescribed both a GLP-1 RA and lithium versus those prescribed a GLP-1 RA only.

A negative control outcome consisting of benign dermatologic conditions unrelated to renal injury was also assessed, including melanocytic nevi (D22.0-D22.9), benign cutaneous neoplasms (D23.0-D23.9), and seborrheic keratoses (L82.0-L82.1). Follow-up extended from the index date until the earliest of a primary outcome event, negative control event, death, loss to follow-up, or completion of the 2-year follow-up period.

### Covariates

Baseline covariates included age, sex, hypertension, disorders of lipoprotein metabolism, overweight and obesity, and ischaemic heart disease. All covariates were identified from diagnoses recorded during the defined exposure interval preceding the index date.

### Propensity score matching

Propensity scores were estimated using logistic regression including all baseline covariates. Exposed and comparator patients were matched one to one using nearest-neighbour matching without replacement and a caliper of 0·1 of the pooled standard deviation of the logit of the propensity score. Covariate balance was assessed using standardized differences, with values less than 0·1 indicating adequate balance. Matching was performed independently for each analytic window and for each of the four sensitivity comparisons.

### Statistical analysis

Event counts, risks, risk differences, and risk ratios were calculated from the matched cohorts. Kaplan-Meier methods and log-rank tests were used for time-to-event analysis. Cox proportional hazards models generated hazard ratios with 95% confidence intervals. All statistical procedures were conducted using the validated TriNetX analytics engine. A two-sided alpha of 0.05 indicated statistical significance.

## Results

### Study population (Figure 1)

A total of 462 matched patient pairs were included in the comparison of diabetes patients treated with lithium plus a GLP-1 RA versus lithium alone. Propensity-score matching using the TriNetX 1:1 nearest-neighbour algorithm achieved excellent covariate balance, with all standardized differences <0.05.

**Figure 1.**
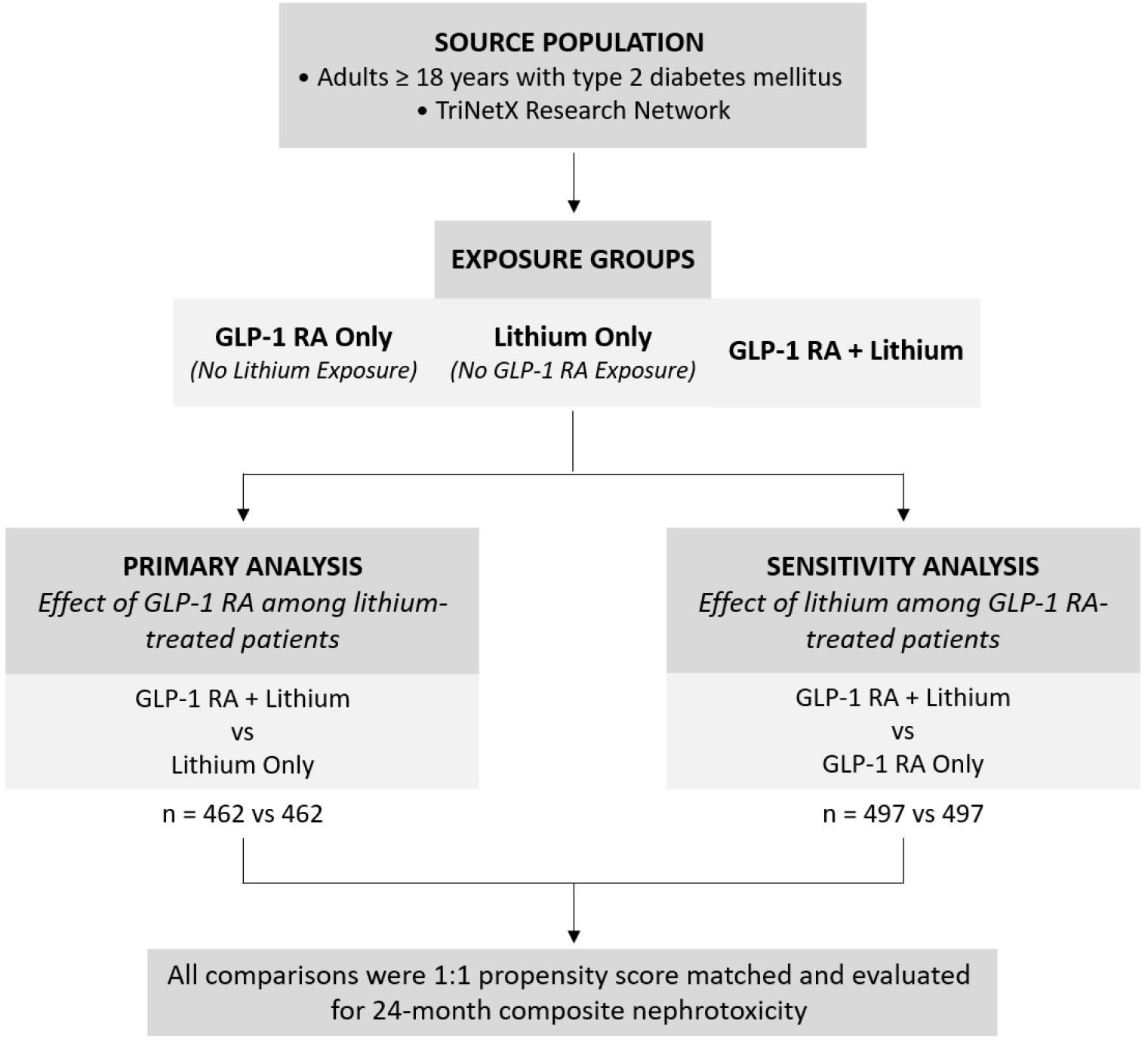
Patient selection flowchart and formation of study comparator groups using pairwise matched contrasts to isolate the independent associations of GLP-1 receptor agonists and lithium.

The cohorts were closely aligned in age (48·7±13·2 vs 48·5±14·0 years; SDiff=0·02) and had identical sex distribution (women 69·3% vs 69·3%; SDiff<0·0001) **(Table 1 & S1)**. Cardiometabolic comorbidities were also well balanced, including T2DM (24·7% vs 25·1%), overweight or obesity (19·5% vs 17·7%), hypertension (11·9% vs 10·6%), dyslipidaemia (10·4% vs 9·5%), and ischaemic heart disease (2·2% vs 2·2%), with standardized differences ranging from <0·0001 to 0·05.

**Table 1.** Baseline characteristics of adults with type 2 diabetes mellitus who received lithium in the prior 24 months, comparing GLP-1 receptor agonist initiators with lithium-treated controls unexposed to GLP-1 receptor agonists, before and after propensity score matching.

|  | Before propensity score matching |  |  | After propensity score matching |  |  |
| --- | --- | --- | --- | --- | --- | --- |
|  | GLP-1 RA-exposed lithium cohort | GLP-1 RA-naïve lithium cohort | SMD | GLP-1 RA-exposed lithium cohort | GLP-1 RA-naïve lithium cohort | SMD |
| Total number | 486 | 2366 | - | 448 | 448 | - |
| Age at the index event, years, mean ± s.d. | 48.6 ± 13.1 | 52.4 ± 15.6 | 0.27 | 48.7 ± 13.2 | 48.5 ± 14 | 0.016 |
| Sex (%) |  |  |  |  |  |  |
| Female | 70.9% | 57.6% | 0.28 | 69.3% | 69.3% | < 0.000 |
| Male | 28.9% | 42.4% | 0.28 | 30.5% | 30.5% | < 0.000 |
| Type 2 Diabetes Mellitus | 27.9% | 10.8% | 0.44 | 24.7% | 25.1% | 0.010 |
| Overweight and Obesity | 23.7% | 5.1% | 0.55 | 19.5% | 17.7% | 0.045 |
| Hypertensive Diseases | 11.2% | 12.5% | 0.040 | 11.9% | 10.6% | 0.041 |
| Disorders of Lipoprotein Metabolism and Other Lipidemias | 10.4% | 9.0% | 0.049 | 10.4% | 9.5% | 0.029 |
| Ischaemic Heart Diseases | 2.0% | 2.9% | 0.060 | 2.2% | 2.2% | < 0.000 |

Follow-up time (i.e., duration of GLP-1 RA exposure) was comparable between groups (mean 293·9 vs 340·5 days; median 291 vs 353·5 days; IQR 344 vs 447 days). Each sensitivity comparison used an independently matched cohort drawn from the broader diabetes population.

### Primary analysis: renal outcomes in persons with T2DM prescribed lithium and a GLP-1RA versus persons with T2DM prescribed lithium and no GLP-1 RA (Figure 2)

For participants who were prescribed lithium within at least 24 months prior to a GLP-1 RA prescription, a lower incidence of renal outcomes compared with those treated with lithium alone was observed (6·1% vs 10·4%; risk difference -4·3% [95% CI -7·86 to -0·80]; risk ratio 0·58 [95% CI 0·37-0.91]; p=0·017). Kaplan-Meier curves revealed consistent time-to-event findings with 24-month event-free survival of 89·0% versus 83·2% (log-rank p=0·037). Overall, these findings indicate that adding a GLP-1 RA to chronic lithium therapy corresponds to a 4.3 percent absolute and 42 percent relative reduction in renal events, reflecting a clinically meaningful improvement over lithium therapy without a GLP-1 RA.

**Figure 2.**
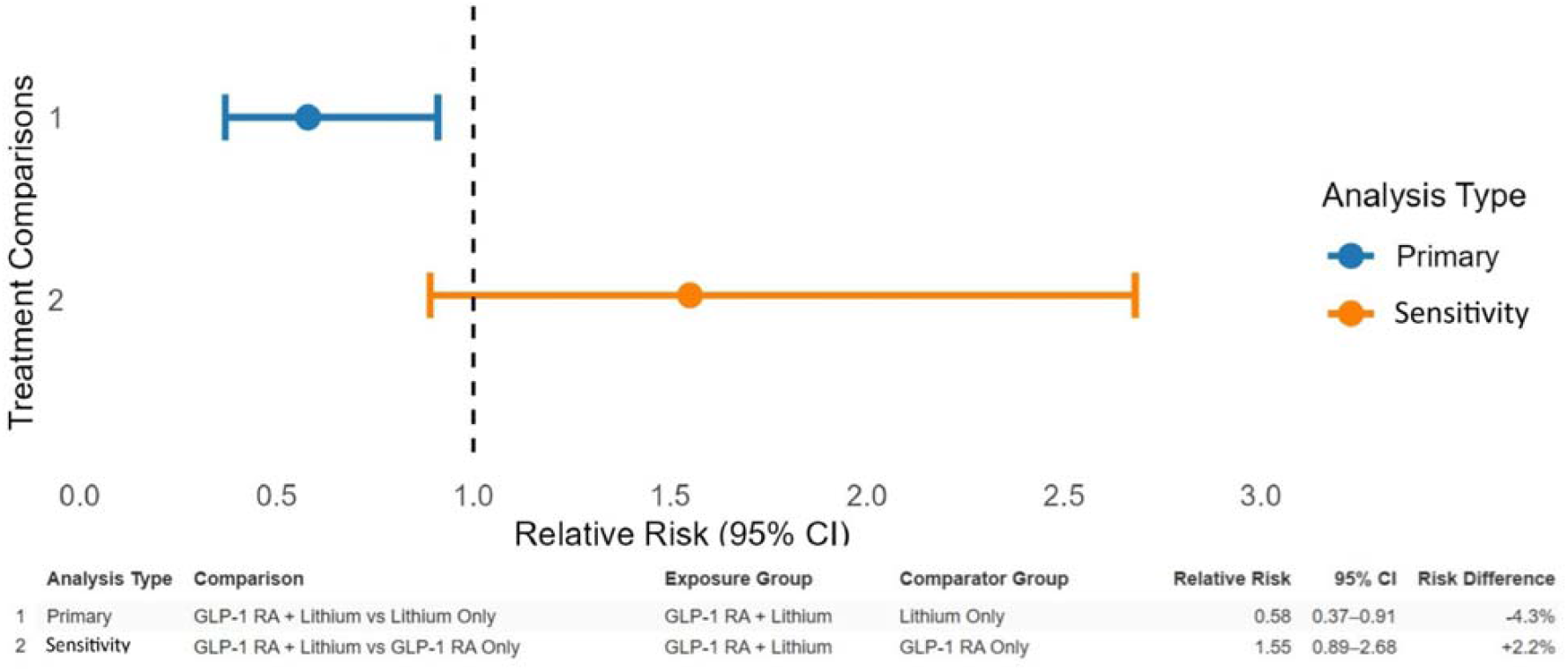
Relative risks of composite nephrotoxicity across primary and sensitivity matched treatment comparisons in adults with type 2 diabetes. The forest plot displays relative risks and 95% confidence intervals from independent 1:1 propensity score– matched cohorts. The primary analysis *(blue)* compares adults with type 2 diabetes and prior lithium exposure who initiated a GLP-1 receptor agonist with matched lithium-treated patients unexposed to GLP-1 receptor agonists. The sensitivity analysis *(orange)* compares adults treated with GLP-1 receptor agonists plus lithium with matched adults treated with GLP-1 receptor agonists alone.

### Sensitivity analysis: persons with T2DM prescribed a GLP-1 RA and lithium versus persons with T2DM prescribed a GLP-1 RA only

Among 497 matched patients per group, GLP-1 RA plus lithium showed a numerically higher 24-month renal event rate than GLP-1 RA therapy alone (6·2% vs 4·0%; risk difference -2·2% [95% CI -0·53 to 4·95]; risk ratio 1·55 [95% CI 0·89-2·68]; p=0·1138). This difference was not statistically significant, and confidence intervals crossed the null. Kaplan-Meier curves showed visible separation (93·0% vs 88·9%; log-rank p<0·0001). While the point estimate suggests a possible 55 percent higher risk when lithium is added, the wide confidence intervals and lack of significance indicate that no reliable difference can be concluded from these data.

### Negative control analysis

Among 462 matched patient pairs, benign dermatologic outcomes occurred at similar rates between exposure groups (3·9 percent versus 2·2 percent; risk difference +1·7 percent [95 percent CI -0·48 to 3·94]; risk ratio 1·80 [95 percent CI 0·84-3·86]; p = 0·13). Kaplan–Meier curves showed no meaningful separation, with 24-month event-free survival remaining comparable across groups.

## Discussion

In this large multi-institutional cohort of adults with T2DM receiving lithium therapy, initiating a GLP-1 RA lowered renal risk by approximately 40% in persons who have taken lithium at least once in the past two years. Results from our sensitivity analysis were in accordance with our primary finding insofar as GLP-1 RA prescription in persons with T2DM also receiving lithium resulted in a risk of lithium-associated nephrotoxicity not significantly different from persons with T2DM receiving GLP-1 RA only.

The nephroprotective effects of GLP-1 RAs in T2DM are not fully accounted for by improvement in metabolic parameters alone. For example, GLP-1 RAs are reported to lower albuminuria by 20 to 40% and slow eGFR decline across multiple independent studies and meta-analyses.^14,22–27^ Experimental and translational studies have reported GLP-1 receptor signaling in renal vascular and tubular cells wherein they suppress inflammatory and oxidative pathways, reducing fibrotic injury.^28,29^ GLP-1 receptor agonists enhance tubular and microvascular resilience via multiple pathways including enhanced renal perfusion, reduced hyperfiltration, and natriuresis with associated reduced blood-pressure. In addition, GLP-1 RAs also enhance lipid disposition, reducing measures of lipotoxic stress, which may also be nephroprotective.^16,25,28^ Results from the FLOW study reported that semaglutide reduced major kidney events by 24% and reduced annual eGFR loss; results that have been reinforced by separate meta-analyses that have documented 16 to 19% reductions in kidney failure and related outcomes.^14,23–25^ In contrast, non-GLP-1 antidiabetic therapies provide glycemic control and produce modest renal benefit.^27,30^ Interpreting our results calls attention to the distinct pathophysiology of lithium-induced nephropathy, which overlap with but is not identical to mechanisms implicated in diabetic kidney disease.^31–33^ For example, chronic lithium therapy results in a predominantly tubulointerstitial pattern of injury with relative glomerular sparing.^31,34^ Long-term lithium exposure have been reported to result in chronic tubulointerstitial nephritis, tubular atrophy, interstitial fibrosis, and numerous 1 to 2 millimeter microcysts in distal nephron segments.^31,32^ Renal microcysts are a defining feature of lithium-induced nephrotoxicity, alongside biopsy and autopsy study reports of patchy scarring with tubular dilatation affecting up to 62% of long-term lithium users.^32,35^ Histopathology data suggest that significant injury typically appears after long-term exposure and would not be expected with acute treatment alone.^32,36^

### Strengths and Limitations

The strength of our analysis is that, to our knowledge, this is the largest cohort of individuals with lithium exposure within the preceding 24 months and were also prescribed a GLP-1 RA, and the overarching research question is whether GLP-1 RAs reduce composite nephrotoxicity risk in this context.^20^ Use of the TriNetX platform and PSM enhances confidence in the findings by balancing key covariates, reducing bias inherent to spontaneous reporting strengthening confidence in our findings. As most individuals receiving lithium have mood disorders, this study also provides the first population-level assessment of renal outcomes with GLP-1 RA therapy in a psychiatric population.

The limitations arise from the retrospective observational design. Although we used PSM, we cannot rule out the possibility of residual confounding. Unmeasured factors relevant to renal vulnerability, including cumulative lithium exposure, serum levels, treatment duration, concomitant nephrotoxins, and baseline tubular function, may also influence the observed effect estimates.^37^ In addition, we categorized GLP-1 RAs as a single class of agents recognizing their pharmacological heterogeneity and also our outcome measure was a composite renal outcome which prevents any conclusions about specific renal disease(s) that are potentially mitigated. We also recognize that our lithium exposure interval is primarily focused on prescription over the last 24 months, a window that we consider relatively brief when evaluating lithium-associated nephrotoxicity. Moreover, we cannot precisely estimate how long people have been receiving lithium or fidelity to taking either lithium or GLP-1 RA. In addition, we cannot completely exclude the possibility of immortal time bias, as nephrotoxicity related to prior lithium exposure may have predated the index exposure date but not have been fully captured in structured electronic health record data. Moreover, we compared lithium-exposed persons with GLP-1 RA exposure to those lithium-exposed without GLP-1 RA exposure, which may introduce the potential for forward-looking exposure classification and related bias, as treatment status in observational data may be influenced by clinical factors and treatment decisions occurring after cohort entry that are not fully captured in the dataset. Furthermore, because outcomes and exposures were identified using coded electronic health record data, misclassification of the timing or occurrence of renal events cannot be excluded, and some renal outcomes may have represented prevalent rather than strictly incident events. Residual confounding related to treatment selection and differences in underlying clinical risk between exposure groups may also persist despite analytic adjustment, and the negative control outcome may not fully share the same confounding structure as renal injury.

Our methodology does not exclude the possibility that the GLP-1 RA mediated nephroprotective effects are not specific to lithium but represent a general renal protective effect. It is notable however that the preliminary evidence does suggest that GLP-1 RAs potentially mitigate renal toxicity from well known nephrotoxic agents (e.g., cisplatin, tacrolimus), suggesting that GLP-1 RAs are targeting molecular and cellular systems relevant to nephrotoxicity from multiple causes including iatrogenic.^18,19,38–40^

## Conclusion

Results from our study provide preliminary support for the hypothesis that GLP-1 RAs may have protective effects against lithium-associated nephrotoxicity. We did not address the pharmacokinetic interactions between lithium and GLP-1 RA exposure, which has been inadequately characterized henceforth in the literature. Case reports of lithium-related toxicity in persons exposed to incretin agonists have appeared, which have been attributed to reductions in volume status due to dehydration, polyuria, and/or vomiting, and/or effects on absorption due to delayed gastric emptying and/or displacement of lithium from the muscle compartment due to muscle wasting.^41,42^ Replication, observational, clinical, as well as mechanistic studies are warranted to further strengthen confidence in the findings. More specifically, whether GLP-1 receptor agonists alleviate histopathologic evidence of lithium-associated nephrotoxicity should be a priority research consideration.

## Data Availability

The individual-level electronic health record data used in this study are not publicly available because access is restricted by contractual agreements and institutional data use policies of the TriNetX research network. Researchers may obtain access to the data through TriNetX subject to institutional approval and a data use agreement. Aggregate results are provided in the manuscript or available from the authors upon reasonable request.

## Acknowledgements

None. The authors received no payments, support, or editorial assistance from pharmaceutical companies, commercial entities, or external agencies. No medical writer or external editor contributed to this manuscript; all writing and revisions were performed solely by the listed authors. All authors had full and unrestricted access to the data and take complete responsibility for its accuracy, integrity, and the decision to submit for publication.

## Author Contributions

Roger S. McIntyre and Angela T.H. Kwan had full access to all TriNetX study data, verified the underlying data, and take responsibility for the integrity and accuracy of the analyses. Both authors serve as guarantors of the study, accepting full responsibility for the overall integrity of the work, with full access to the data, and final authority over the decision to publish.

All authors meet ICMJE authorship criteria, contributed to data interpretation, and accept responsibility for the decision to submit the manuscript.

Concept and Design: McIntyre

Acquisition, Analysis, or Interpretation of Data: Kwan Drafting of the Manuscript: McIntyre, Kwan

Critical Revision of the Manuscript for Important Intellectual Content: All authors Statistical Analysis: Kwan

Administrative, Technical, or Material Support: McIntyre Supervision: McIntyre

## Ethical Considerations

This study was conducted using fully deidentified electronic health record data from the TriNetX Research Network. Institutional Review Board approval was obtained at SUNY Upstate Medical University (IRB #2302956-2). Because TriNetX contains only deidentified patient information, informed consent was not required.

