## Supplemental Table 1 for "Nephroprotective Effect of GLP-1 Receptor Agonists (GLP-1 RAs) in Patients Receiving Lithium Therapy: A Population-Based Study Using the TriNetX Network"

**Supplementary Table**

**Table S1.** Baseline characteristics of adults with type 2 diabetes mellitus with and without GLP-1 receptor agonist exposure, before and after propensity score matching.

|  | **Before propensity score matching** | | |  | **After propensity score matching** | | |
| --- | --- | --- | --- | --- | --- | --- | --- |
|  | **GLP-1 RA-exposed cohort** | **T2DM cohort** | **SMD** |  | **GLP-1 RA-exposed cohort** | **T2DM cohort** | **SMD** |
| Total number | 228,156 | 9,498,482 | - |  | 94,564 | 94,564 | - |
| Age at the index event, years, mean ± s.d. | 56·9 ± 12·9 | 60·3 ± 15·2 | 0·2393 |  | 56·8 ± 12·9 | 56·8 ± 12·9 | < 0.0001 |
| Sex (%) |  |  |  |  |  |  |  |
| Female | 65·44% | 50·00% | 0·3165 |  | 63·12% | 63·12% | < 0.0001 |
| Male | 34·49% | 49·93% | 0·3165 |  | 36·87% | 36·87% | < 0.0001 |
| Type 2 Diabetes Mellitus | 41·45% | 100% | 1·6809 |  | 100% | 100% | - |
| Overweight and Obesity | 22·23% | 12·91% | 0·25 |  | 31·80% | 31·80% | < 0.0001 |
| Hypertensive Diseases | 22·51% | 51·77% | 0·64 |  | 41·85% | 41·85% | < 0.0001 |
| Disorders of Lipoprotein Metabolism and Other Lipidemias | 19·76% | 35·12% | 0·35 |  | 37·87% | 37·87% | < 0.0001 |
| Ischaemic Heart Diseases | 2·95% | 13·05% | 0·38 |  | 4·71% | 4·71% | < 0.0001 |
